# A joint transcriptome-wide association study across multiple tissues identifies new candidate susceptibility genes for breast cancer

**DOI:** 10.1101/2022.09.30.22280575

**Authors:** Guimin Gao, Peter N. Fiorica, Julian McClellan, Alvaro Barbeira, James L. Li, Olufunmilayo I. Olopade, Hae Kyung Im, Dezheng Huo

**Affiliations:** Department of Public Health Sciences, University of Chicago, IL, 60637, USA; Section of Genetic Medicine, Department of Medicine, University of Chicago, IL, 60637, USA; Section of Hematology & Oncology, Department of Medicine, University of Chicago, IL, 60637, USA

## Abstract

Genome-wide association studies (GWAS) have identified more than 200 genomic loci for breast cancer risk, but specific causal genes in most of these loci have not been identified. In fact, transcriptome-wide association studies (TWAS) of breast cancer performed using gene expression prediction models trained in breast tissue have yet to clearly identify most target genes. To identify novel candidate genes, we performed a joint TWAS analysis that combined TWAS signals from multiple tissues. We used expression prediction models trained in 47 tissues from the Genotype-Tissue Expression data using a multivariate adaptive shrinkage method along with association summary statistics from the Breast Cancer Association Consortium and UK Biobank data. We identified 380 genes at 129 genomic loci to be significantly associated with breast cancer at the Bonferroni threshold (p < 2.36 × 10^−6^). Of them, 29 genes were located in 11 novel regions that were at least 1Mb away from published GWAS hits. The rest of TWAS-significant genes were located in 118 known genomic loci from previous GWAS of breast cancer. After conditioning on previous GWAS index variants, we found that 22 genes located in known GWAS loci remained statistically significant. Our study maps potential target genes in more than half of known GWAS loci and discovers multiple new loci, providing new insights into breast cancer genetics.

## Introduction

Breast cancer is the most common malignancy among women in most countries around the world, accounting for one quarter of all cancer cases in women^1^. In the past fifteen years, genome-wide association studies (GWAS) have identified over 200 loci significantly associated with breast cancer ^2-4^. While some of these findings have yielded functional insights into breast cancer ^4^, these genetic variants account for a relatively small proportion of heritability, suggesting more genetic variants have yet to be identified. Because the vast majority of risk variants identified in GWAS are located in intergenic regions and are not nonsynonymous coding variants, the putative genes on which these risk variants act to cause breast cancer remain unclear for most GWAS-identified loci.

To further elucidate the role of genetic variants in complex traits, transcriptome-wide association studies (TWAS) have been conducted to quantify the relationship between a predicted level of genetically regulated gene expression and the phenotype of interest ^5,6^. TWAS of breast cancer have identified dozens of genes whose expression are significantly associated with breast cancer and its subtypes ^4,7-9^. However, these genes only account for only a small proportion of known GWAS loci of breast cancer. These TWAS were performed primarily by associating a cis-regulated level of gene expression with breast cancer in single tissues (breast tissue or whole blood). Our recent study demonstrated that integrating information from multiple tissues in TWAS could improve association detection ^10^. In addition, existing TWASs used gene expression prediction models trained in data from older versions of the Genotype-Tissue expression (GTEx) project, such as versions 6 or 7. The recent version 8 of GTEx has much larger sample sizes compared to the older versions, so the expression models trained in GTEx v8 will be more accurate to predict expression levels than those trained in older versions ^11,12^. By using GTEx v8, one can explore heritability for expression more efficiently and more genes can pass the filtering threshold and be used for TWAS analysis ^12^. Therefore, the prediction models trained in GTEx (v8) have the potential to increase the power of TWAS in detecting susceptibility genes.

In this study, we aimed to identify novel candidate genes for breast cancer by performing joint TWAS analyses of breast cancer by combining TWAS information from multiple tissues. We applied our TWAS method to the summary statistics from a meta-analysis of data from 122,977 breast cancer cases and 105,974 controls in the Breast Cancer Association Consortium (BCAC) ^3^ and 10,534 breast cancer cases and 185,116 controls in UK Biobank (UKB) ^13^.

## Results

### Joint TWAS combining information across multiple tissues

We used expression prediction models trained in 47 tissues of European ancestry (with sample sizes ranging from 65 to 588 and a median of 194) from the GTEx v8 data using a multivariate adaptive shrinkage (MASH) method ^11,12,14^. In total, 21,227 genes across the 47 tissues with prediction models, including 14,634 genes expressed in breast tissue, were tested in our TWAS analysis.

To acquire higher power in the TWAS analysis, we first performed a GWAS analysis in a breast cancer dataset from UKB and then combined the UKB GWAS results with the previously published GWAS results of the BCAC data ^3^ by a meta-analysis with the software METAL^15^. Using meta-analysis summary statistics, we performed a traditional TWAS individually in each tissue by S-Predixcan ^16^, and then joint TWAS using the aggregated Cauchy association test (ACAT) ^17^, which combined p-values of single-tissue TWAS across the 47 tissues.

The results of the joint TWAS analysis are summarized in the Manhattan plots against the variant-based GWAS analysis results (Supplementary Figure S1). Of the 21,227 genes tested in our joint TWAS analysis, we identified 380 genes whose predicted expression was associated with breast cancer risk at the Bonferroni-corrected threshold of p < 2.4×10^−6^ (Supplementary Table S1). Only 141 genes were identified when TWAS analysis used only breast tissue, i.e. conventional single-tissue TWAS approach ^16^ (Supplementary Table S2). Of these 141 genes, 127 genes were also identified in the joint, multi-tissue TWAS. The remaining 14 genes identified only in the breast-tissue TWAS analysis were only marginally significant, so we focused on results from the joint TWAS. Supplementary Table S3 shows the detailed single-tissue TWAS results for the 380 genes in the analysis of two databases (BCAC, UKB) pooled and separately. We found that Z scores across tissues were moderately concordant on average, with an intraclass correlation coefficient of 0.528, but the agreement between tissues as well as the strongest association signals varied across genes. These findings suggest that the multi-tissue joint TWAS could provide additional information compared to traditional single target tissue TWAS, and address the possibility that for different genes the target tissue(s) could vary. We also found that TWAS results using the BCAC and UKB databases were very consistent with a Pearson r = 0.859 (Supplementary Figure S2).

Of the 380 genes identified in our joint TWAS, 32 genes have been reported in previous TWAS (Supplementary Tables 1 and 4), 80 genes have been implicated in previous GWAS, such as *FGFR2, TOX3*, and *ESR1* (Supplementary Tables 1 and 5), and eight genes were reported both in previous TWAS and GWAS, such as *TNFSF10*. The genes implicated by multiple previous GWAS studies were more likely to be re-discovered in our TWAS. The 380 genes are distributed among 129 genomic loci (Figure 1). Based on NHGRI-EBI GWAS Catalog ^18^ and literature review, we curated 222 GWAS loci of breast cancer susceptibility (Supplementary Table 6). Our joint TWAS identified 351 significant genes that are located in 118 known GWAS susceptibility loci. The remaining 29 genes are located in 11 novel loci that are at least 1Mb away from any risk variant identified in previous GWAS and are not in linkage disequilibrium (LD) with risk variants (Table 1). Of the 29 genes found in novel loci, 13 genes in 7 loci were also significant in the breast tissue-based TWAS at the Bonferroni threshold. For example, we found *MAP2K4* in the 17p12 locus was significant in both multi-tissue joint TWAS and breast tissue-based TWAS, although there was no reported GWAS signal in this locus (Figure 2).

**Figure 1.**
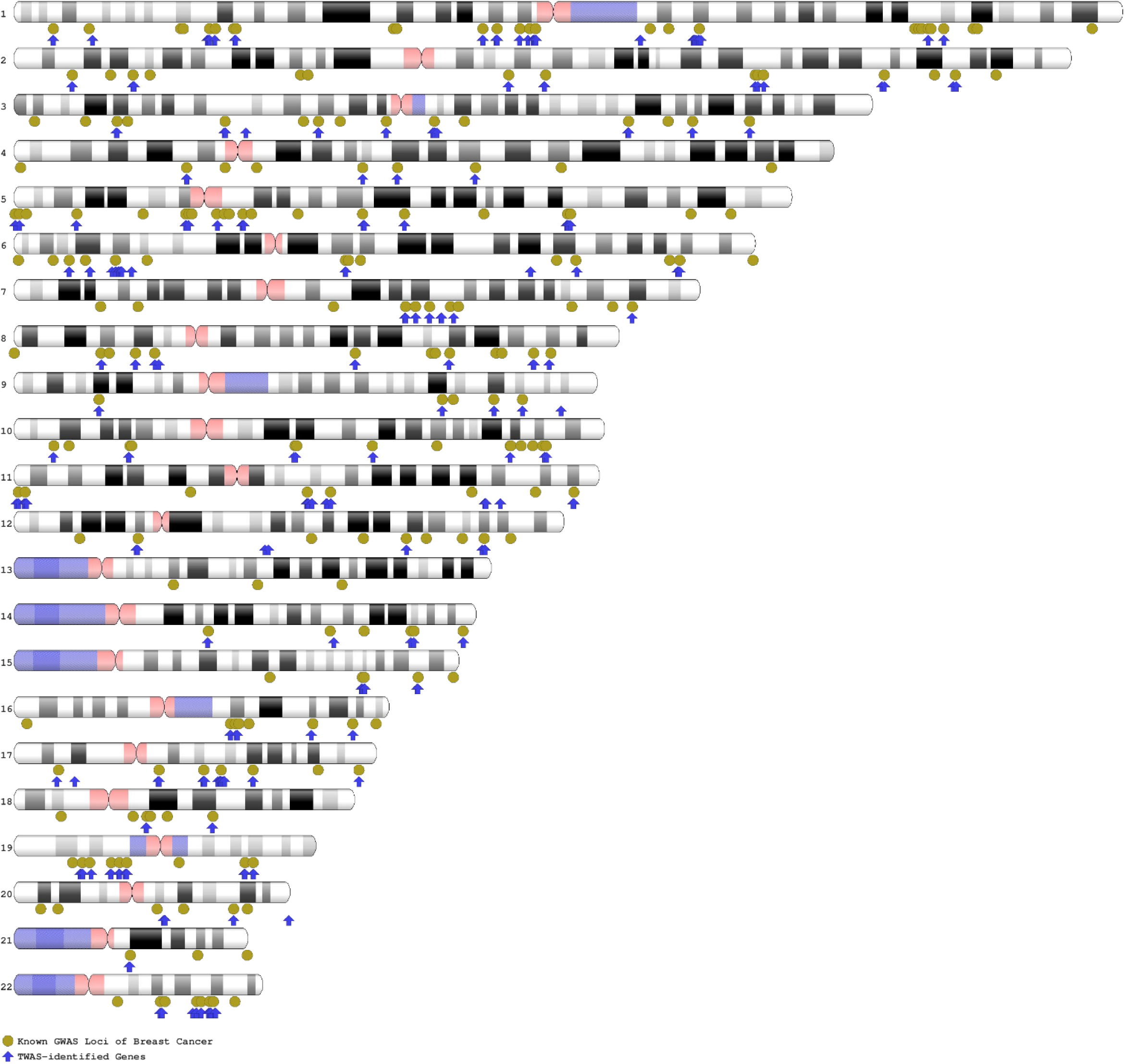
Ideogram of the 380 genes in the context of known GWAS loci of breast cancer.

**Figure 2.**
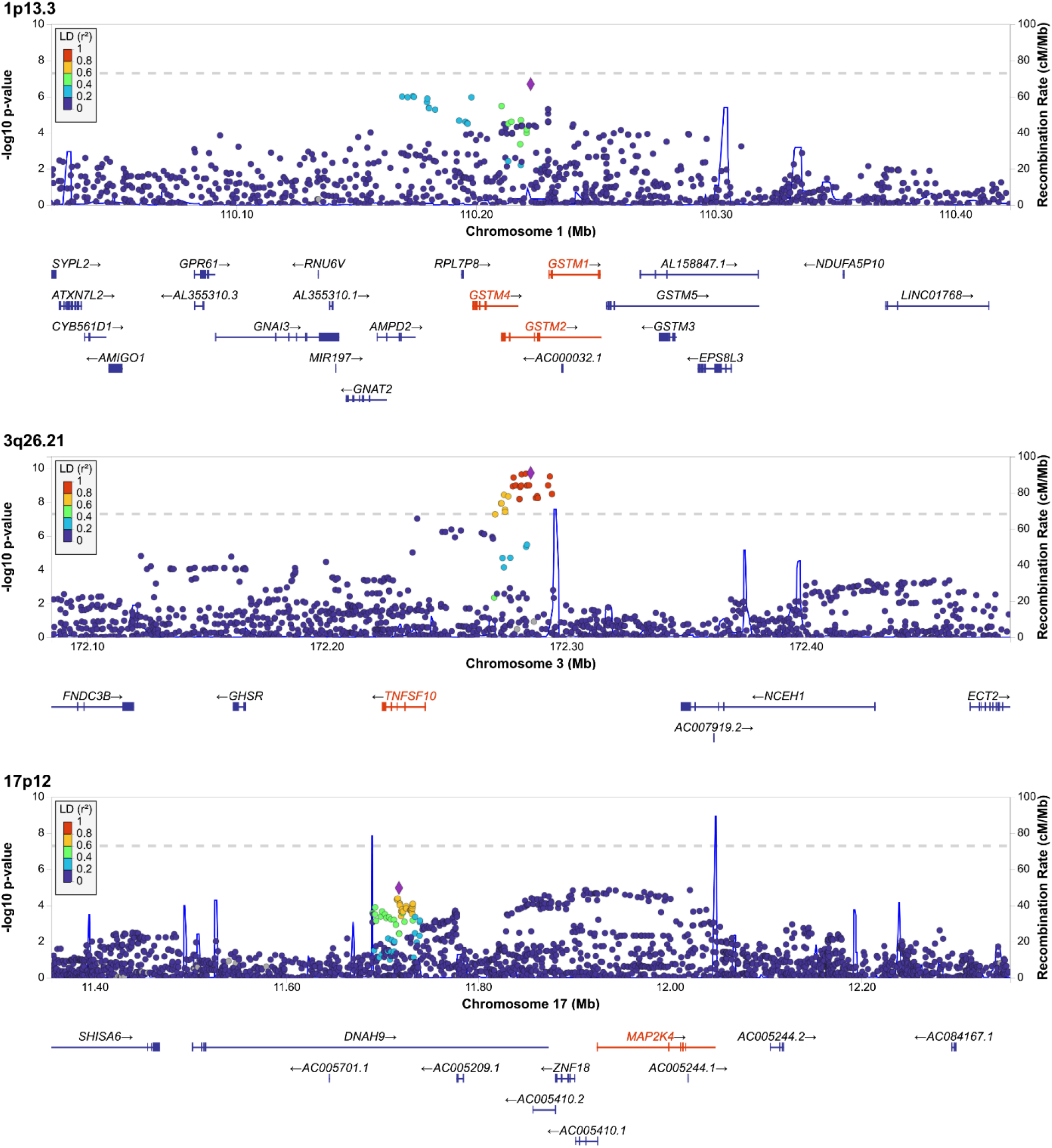
Exemplar genes in 3 loci identified by the joint TWAS analysis.

**Table 1.**
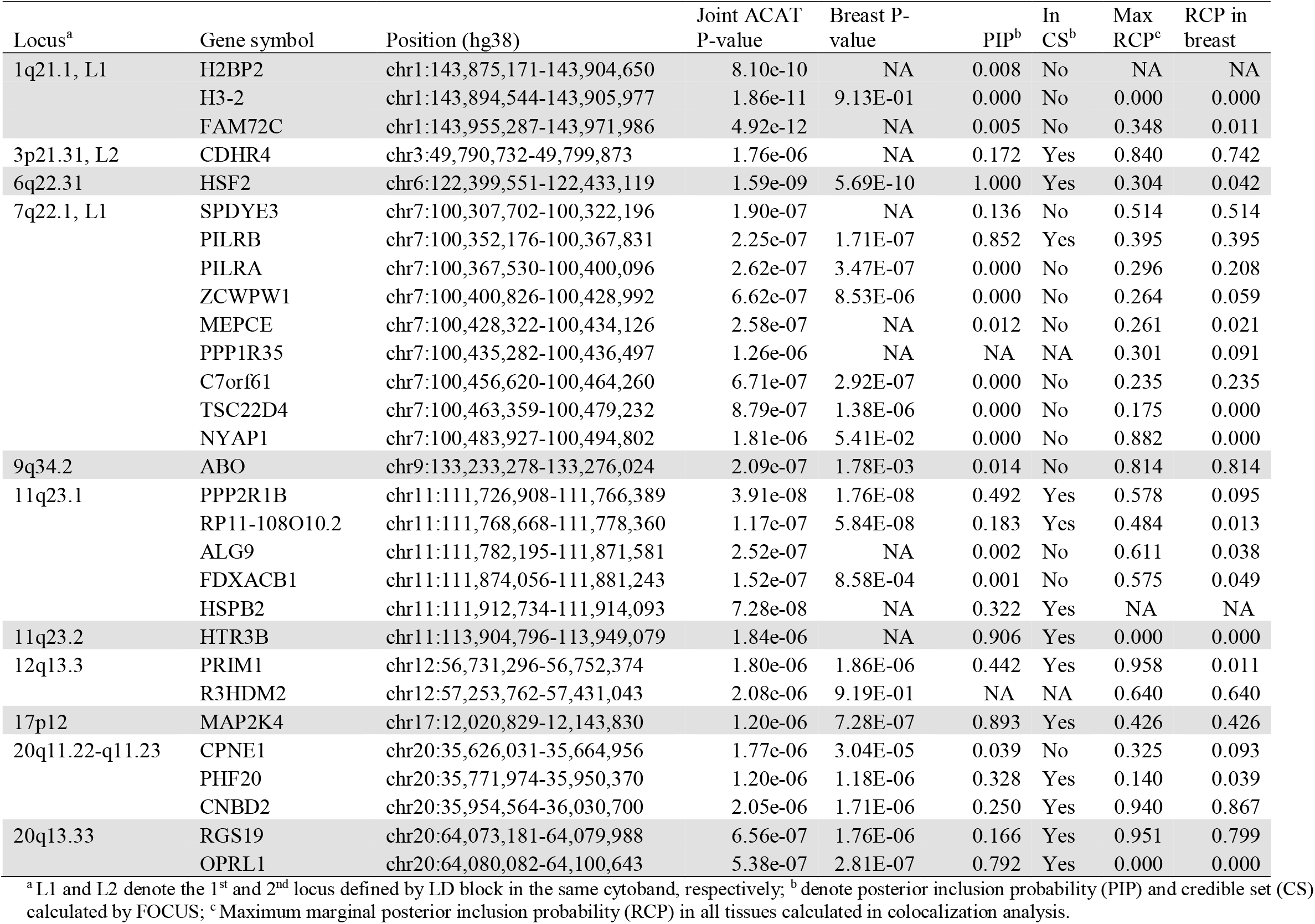
The 29 genes identified by joint TWAS located at 11 genomic loci at least 1 Mb away from previous GWAS hits.

### Conditional joint TWAS on published GWAS index variants

To determine whether the associations for the genes identified by the joint TWAS were independent of GWAS association signals, we performed conditional analyses adjusting for nearby GWAS index risk variants. We found 22 genes located in 12 known GWAS loci that were conditionally significant (Table 2). This suggests that additional genetic variants, which are neither genome-wide significant nor in LD with GWAS significant variants, may account for the association between expression of these genes and breast cancer risk at these loci.

**Table 2.**
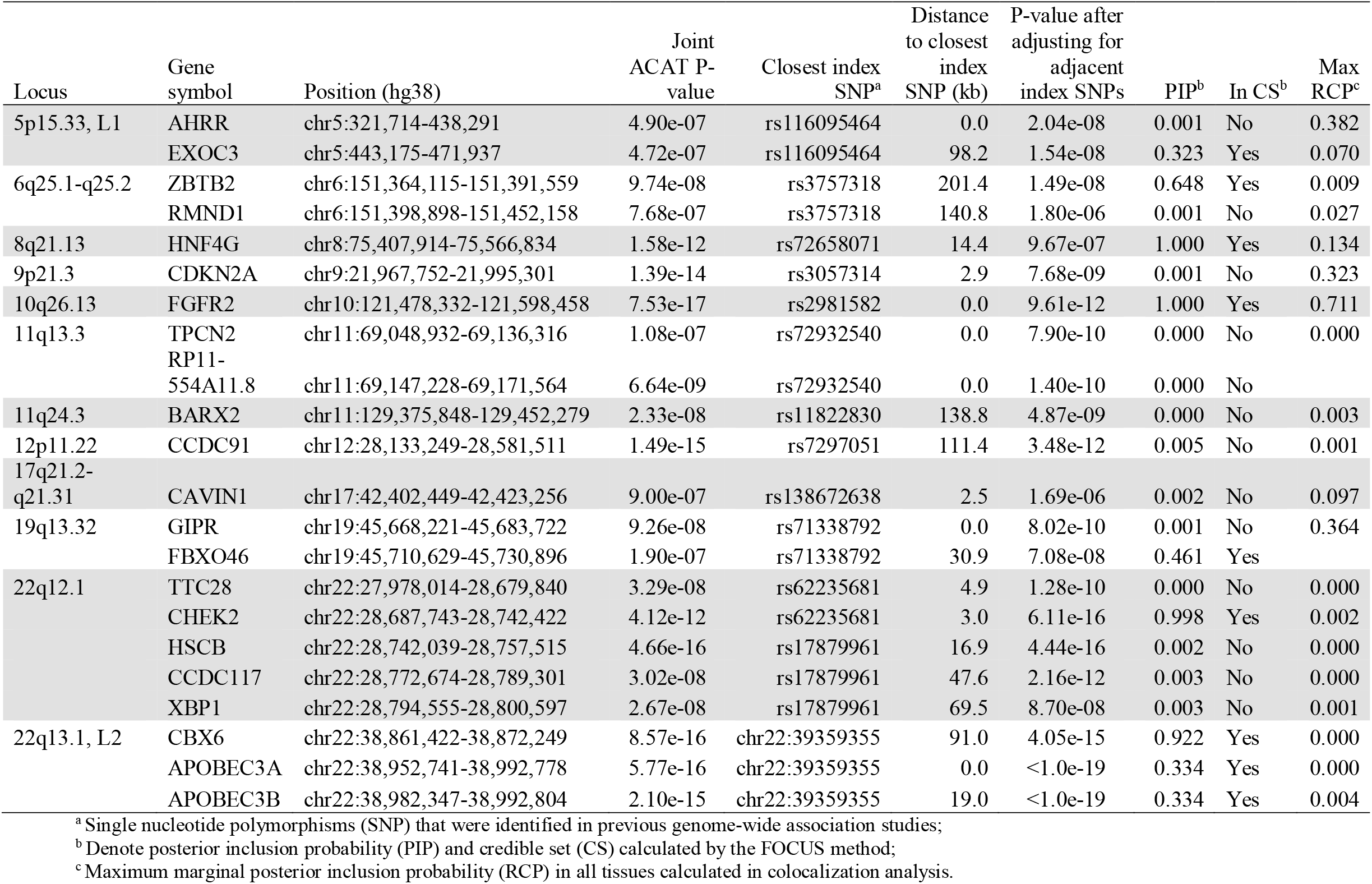
The 22 genes identified by joint TWAS in the 12 known loci and significant after adjusting for known GWAS SNPs.

### Colocalization analysis and gene-based fine mapping

Within an LD block region, the LD among single nucleoid polymorphisms (SNPs) can induce significant gene-trait associations for non-causal genes as a function of the expression quantitative trait loci (eQTL) weights used in predicting expression ^19^. To identify causal genes among the 380 TWAS-identified genes, we performed colocalization analysis and gene-based fine mapping. For colocalization, we used the package ENLOC ^20^ to identify evidence of colocalization between the GWAS and the eQTL signals by calculating regional colocalization probabilities (RCP). An RCP at a gene greater than a threshold (such as 0.5) provides supportive information that the gene identified by the joint TWAS has a high probability of being causal. In our analysis, 110 of 380 genes had RCP values greater than 0.5 (Supplementary Table 1).

In the gene-based fine-mapping of TWAS using the package FOCUS ^19^, we generated credible sets of genes at the confidence level of 90% and calculated marginal posterior inclusion probability (PIP) for each gene. If a gene is in a credible set and has a high PIP, then the gene is likely to be causal. In our fine-mapping analysis, 140 genes were found to be in credible sets.

Based on the co-localization and fine mapping analyses, there were 58 genes in credible sets and with RCP values greater than 0.5, exhibiting strong evidence of being causal genes (Table 3). These 58 genes were located in 47 loci. For most loci, co-localization and fine-mapping identified only one causal gene candidate, eliminating many TWAS-identified genes; for example, *ASH1L* in locus 1q22 (out of 18 TWAS-identified genes) and *FGFR2* in locus 10q26.13 (out of 3 TWAS-identified genes) were identified as possible causal genes. For fewer loci, multiple candidate genes were identified after co-localization and fine mapping; for example, *GSTM1, GSTM2*, and *GSTM4*, three members of the glutathione S-transferase multigene family, were suggested to be possible causal genes in locus 1p13.3 in our analysis (Table 3, Figure 2).

**Table 3.** The 58 candidate causal genes in 47 loci that are in the credible set and RCP>0.5.

| Locus | Gene symbol | Position (hg38) | Joint ACAT<br>P-value | Breast<br>P-value | PIP<sup>a</sup> | Max<br>RCP<sup>b</sup> | RCP in<br>breast |
| --- | --- | --- | --- | --- | --- | --- | --- |
| 1p36.22 | PEX14 | chr1:10,472,288-10,630,758 | 7.45e-16 | 2.65E-10 | 0.288 | 0.707 | 0.002 |
| 1p36.13 | KLHDC7A | chr1:18,480,930-18,485,974 | 1.51e-15 | 5.19E-13 | 1.000 | 1.000 | 0.038 |
| 1p34.1-1p33 | PIK3R3 | chr1:46,040,140-46,133,036 | 7.22e-08 | 4.16E-08 | 0.848 | 0.800 | 0.062 |
| 1p13.3 | GSTM4 | chr1:109,656,099-109,674,836 | 2.71e-07 | 1.27E-05 | 0.146 | 0.994 | 0.994 |
|  | GSTM2 | chr1:109,668,022-109,709,551 | 2.85e-08 | 8.63E-07 | 0.200 | 0.994 | 0.994 |
|  | GSTM1 | chr1:109,687,814-109,709,039 | 2.28e-09 | 6.79E-09 | 0.495 | 0.995 | 0.995 |
| 1p13.2 | DCLRE1B | chr1:113,905,213-113,914,086 | 3.67e-10 | 2.35E-08 | 0.986 | 0.949 | 0.949 |
| 1q22 | ASH1L | chr1:155,335,268-155,563,162 | 6.06e-11 | 1.52E-11 | 0.879 | 0.888 | 0.888 |
| 1q32.2 | LINC02767 | chr1:207,959,292-207,969,329 | 1.35e-07 | 3.01E-08 | 0.992 | 0.630 | 0.630 |
| 2p23.3 | ADCY3 | chr2:24,819,169-24,920,237 | 2.18e-10 | NA | 0.449 | 0.691 | 0.128 |
| 2q31.1, L2 | DLX2 | chr2:172,099,438-172,102,900 | 3.71e-13 | 1.33E-13 | 1.000 | 0.919 | 0.209 |
| 2q33.1 | CASP8 | chr2:201,233,443-201,361,836 | 8.45e-17 | 1.80E-14 | 0.993 | 0.868 | 0.000 |
| 3p12.1 | VGLL3 | chr3:86,876,388-86,991,149 | 3.00e-10 | 1.20E-10 | 1.000 | 1.000 | 0.865 |
| 3q23 | ZBTB38 | chr3:141,324,213-141,449,792 | 2.75e-16 | 2.04E-17 | 1.000 | 0.764 | 0.710 |
| 4p14 | TLR10 | chr4:38,772,238-38,782,990 | 9.88e-13 | 4.05E-13 | 0.930 | 0.553 | 0.245 |
|  | FAM114A1 | chr4:38,867,677-38,945,739 | 1.76e-10 | 3.81E-05 | 0.263 | 0.866 | 0.866 |
| 4q21.23 | MRPS18C | chr4:83,455,932-83,469,735 | 2.06e-07 | 5.47E-05 | 0.180 | 0.574 | 0.034 |
| 4q22.1 | PPM1K | chr4:88,257,620-88,284,769 | 4.62e-08 | 1.03E-09 | 1.000 | 0.970 | 0.970 |
| 5p15.1 | MARCHF11 | chr5:16,067,139-16,180,762 | 9.36e-13 | 7.80E-14 | 1.000 | 0.584 | 0.584 |
| 5q14.1-q14.2 | ATG10 | chr5:81,972,023-82,276,857 | 2.79e-13 | 1.71E-08 | 0.389 | 0.738 | 0.738 |
| 5q31.1, L1 | SLC22A5 | chr5:132,369,710-132,395,613 | 2.08e-10 | 4.22E-10 | 0.072 | 0.830 | 0.172 |
|  | IRF1 | chr5:132,440,440-132,508,719 | 5.57e-11 | NA | 0.928 | 0.680 | 0.051 |
| 5q31.1, L2 | HSPA4 | chr5:133,052,013-133,106,449 | 7.48e-09 | 4.98E-01 | 0.843 | 0.638 | 0.638 |
| 6p23 | RANBP9 | chr6:13,621,498-13,711,835 | 4.51e-13 | 6.33E-05 | 0.352 | 0.721 | 0.016 |
| 6q14.1, L1 | RP11-250B2.5 | chr6:80,466,958-80,469,080 | 8.14e-08 | 3.30E-07 | 0.933 | 0.537 | 0.516 |
| 6q22.31 | HSF2 | chr6:122,399,551-122,433,119 | 1.59e-09 | 5.69E-10 | 1.000 | 0.840 | 0.742 |
| 6q23.1 | L3MBTL3 | chr6:130,013,699-130,141,449 | 6.31e-12 | 4.56E-11 | 1.000 | 0.990 | 0.590 |
| 7q21.3, L2 | BAIAP2L1 | chr7:98,291,650-98,401,090 | 5.08e-08 | 2.51E-07 | 0.812 | 0.836 | 0.836 |
| 7q22.1, L1 | PILRB | chr7:100,352,176-100,367,831 | 2.25e-07 | 1.71E-07 | 0.852 | 0.514 | 0.514 |
| 8p11.23 | RP11-419C23.1 | chr8:37,067,441-37,069,418 | 4.87e-17 | 6.32E-22 | 1.000 | 0.920 | 0.896 |
| 10p15.1 | GDI2 | chr10:5,765,223-5,842,132 | 5.40e-07 | 8.15E-08 | 0.987 | 0.711 | 0.424 |
| 10p12.31 | MLLT10 | chr10:21,524,646-21,743,630 | 1.95e-17 | NA | 1.000 | 0.854 | 0.659 |
| 10q26.13 | FGFR2 | chr10:121,478,332-121,598,458 | 7.53e-17 | NA | 1.000 | 0.711 | 0.065 |
| 11p15.5, L1 | PANO1 | chr11:797,511-799,190 | 1.40e-12 | 4.55E-13 | 0.216 | 0.798 | 0.660 |
|  | PIDD1 | chr11:799,179-809,753 | 2.25e-14 | 5.97E-14 | 0.773 | 0.973 | 0.845 |
| 11p15.5, L2 | PRR33 | chr11:1,888,078-1,891,895 | 6.08e-17 | 5.65E-27 | 0.990 | 0.644 | 0.145 |
| 11q13.1 | OVOL1 | chr11:65,787,063-65,797,214 | 2.83e-12 | 2.14E-10 | 0.983 | 0.679 | 0.171 |
| 11q13.1 | CTSF | chr11:66,563,464-66,568,879 | 7.44e-07 | 1.10E-04 | 0.065 | 0.773 | 0.771 |
| 11q23.1 | PPP2R1B | chr11:111,726,908-111,766,389 | 3.91e-08 | 1.76E-08 | 0.492 | 0.814 | 0.814 |
|  | RP11-108O10.2 | chr11:111,768,668-111,778,360 | 1.17e-07 | 5.84E-08 | 0.183 | 0.578 | 0.095 |
|  | HSPB2 | chr11:111,912,734-111,914,093 | 7.28e-08 | NA | 0.322 | 0.575 | 0.049 |
| 12q22 | NTN4 | chr12:95,657,807-95,791,189 | 1.95e-16 | 2.46E-41 | 1.000 | 0.964 | 0.952 |
| 14q13.3 | PAX9 | chr14:36,657,568-36,679,362 | 4.74e-17 | 5.61E-20 | 1.000 | 0.829 | 0.771 |
| 15q24.1 | ULK3 | chr15:74,836,118-74,843,346 | 3.56e-08 | 6.78E-08 | 0.885 | 0.846 | 0.599 |
| 15q26.1 | RCCD1 | chr15:90,954,881-90,963,125 | 5.71e-16 | 1.04E-15 | 1.000 | 0.857 | 0.851 |
| 16q22.2 | ZNF23 | chr16:71,447,597-71,463,095 | 2.88e-07 | 2.73E-06 | 0.465 | 0.673 | 0.673 |
| 17p12 | MAP2K4 | chr17:12,020,829-12,143,830 | 1.20e-06 | 7.28E-07 | 0.893 | 0.640 | 0.640 |
|  | STXBP4 | chr17:54,968,727-55,173,632 | 7.01e-17 | 9.98E-19 | 1.000 | 0.643 | 0.000 |
| 17q25.3 | CBX8 | chr17:79,792,132-79,801,683 | 1.55e-10 | 8.57E-12 | 1.000 | 0.909 | 0.897 |
| 19p13.13 | RAD23A | chr19:12,945,855-12,953,642 | 1.93e-06 | NA | 0.051 | 0.790 | 0.030 |
|  | LYL1 | chr19:13,099,033-13,103,161 | 4.68e-10 | NA | 0.827 | 0.867 | 0.182 |
| 19p13.11, L1 | ABHD8 | chr19:17,292,131-17,310,236 | 1.18e-07 | 6.66E-05 | 0.701 | 0.712 | 0.191 |
| 19p13.11, L2 | SSBP4 | chr19:18,418,864-18,434,562 | 7.67e-18 | 5.76E-24 | 1.000 | 0.757 | 0.757 |
|  | ISYNA1 | chr19:18,434,388-18,438,167 | 1.85e-17 | 2.54E-14 | 0.108 | 0.629 | 0.036 |
| 19q13.31 | KCNN4 | chr19:43,766,533-43,780,976 | 7.65e-17 | NA | 1.000 | 0.962 | 0.143 |
| 20q13.33 | RGS19 | chr20:64,073,181-64,079,988 | 6.56e-07 | 1.76E-06 | 0.166 | 0.940 | 0.867 |
|  | OPRL1 | chr20:64,080,082-64,100,643 | 5.38e-07 | 2.81E-07 | 0.792 | 0.951 | 0.799 |
| 22q13.1, L1 | MAFF | chr22:38,200,767-38,216,507 | 2.50e-14 | NA | 1.000 | 0.715 | 0.123 |

<sup>a</sup> Posterior inclusion probability (PIP) and credible set (CS) calculated by the FOCUS method;
<sup>b</sup> Maximum marginal posterior inclusion probability (RCP) in all tissues calculated in colocalization analysis.

### Gene Set Enrichment and Functional Annotation

Of the 380 TWAS-identified genes, 321 are protein-coding genes, 54 are long non-coding RNA (lncRNA) genes, and 5 are pseudogenes. We tested the enrichment of this set of protein coding and lncRNA genes against background gene sets from multiple databases using the FUMA software package ^21^. We found these TWAS-identified genes were significantly enriched in several biological pathways, such as the Trail signaling pathway, Fas signaling pathway, apoptosis pathway, biosynthesis, and cell cycle regulation; all of these pathways are important in cancer development or a hallmark of cancer, which further warrants efforts into studying how the genes identified in our TWAS may contribute to breast cancer etiology (Supplementary Table S7). Interestingly, we found significant enrichment in the genes underlying several breast cancer risk factors, including body fat distribution, mammographic density, alcohol use, and body height^22^, suggesting that the TWAS-identified genes may indirectly contribute to breast cancer susceptibility through their impacts on known lifestyle/environmental risk factors. We also found strong enrichment for other diseases, such as inflammatory bowel disease, diabetes, and other cancers (including melanoma, chronic myeloid leukemia, and cancers of the esophagus, prostate, and bladder), suggesting that some of breast cancer genes have pleiotropic effects. These results were consistent with the notion that there are shared genetic components between various cancer sites^23^, and suggest that further research into collating TWAS results across cancers may be beneficial to understanding their shared genetic etiologies. Lastly, differential gene expression analysis in GTEx showed that the TWAS-identified genes had strong tissue specificity, although our joint TWAS weighted each tissue similarly; the most up-expressed tissues of these genes were uterus, breast, ovary and vagina (Supplementary Figure S3).

## Discussion

In this study, we performed a breast cancer TWAS analysis that leveraged the genetically predicted gene expression levels across multiple tissues. We identified 380 significant genes at the Bonferroni threshold, including 29 genes located in 11 novel loci and 351 genes located in 118 known GWAS susceptibility loci. In more than half of the known breast cancer GWAS loci, our study was able to identify possible susceptibility gene(s). We also found 22 genes in known GWAS loci that were independent of previously reported GWAS risk variants, suggesting potentially new breast cancer susceptibility signals. Lastly, through co-localization and fine-mapping analysis, we inferred that 58 genes have a high probability of being causal genes. Generally, our study findings are consistent with previous TWASs; of the 78 genes reported in previous TWASs,^4,7-9,24^ 32 genes were replicated in our study.

Our study identified substantially more TWAS significant genes than all previous studies combined, possibly because of several notable differences in methodologies. First, we used GWAS data from a large number of breast cancer cases (N=133,511) and controls (N=291,090) from BCAC and UKB. This large sample size provided high statistical power in the association analysis. If we had used summary statistics exclusively from BCAC like in previous studies ^4,8^, our joint TWAS would have only identified 294 genes as opposed to the 380 genes we identified using both BCAC and UKB datasets. Second, we aggregated TWAS signals across 47 tissues. This multi-tissue approach resulted in more genes being identified compared to the TWAS using breast tissue alone. This suggests that while breast tissue is an important tissue to utilize when conducting breast cancer TWASs, other tissues can contribute additional information for gene discovery. For example, our multi-tissue approach identified *FGFR2*, a gene with strong evidence in breast cancer etiology, but this gene has not been identified in previous TWASs and would have been missed if we had utilized only the TWAS exclusive to breast tissue. Third, we used expression prediction models trained in GTEx v8 with the MASH method based on fine mapping to select possible causal eQTLs as predictors for each gene. The expression models trained in GTEx v8 can be more accurate than those trained in older versions of GTEx for three reasons: a) The sample sizes for tissues in GTEx v8 are larger than those in older versions of GTEx (for example, we used 329 samples from GTEx v8 to build prediction models of breast tissue in European ancestry individuals, while Wu et al. ^8^ used 67 samples from v6); b) Selecting possibly causal eQTL through fine mapping can reduce the probability that non-causal eQTLs were used in the prediction models ^12^; and c) MASH accounts for eQTL correlation across tissues and provides more accurate estimates of beta coefficients of eQTLs used as final weights in the prediction models. By using the prediction models trained in GTEx (v8) data, we were able to perform this joint TWAS analysis on 21,227 genes with prediction models of good performance (i.e., with eQTL signals). In contrast, two previously published large TWAS that relied on breast tissues in older version of GTEx and traditional methods can evaluate a smaller subset of genes^7,8^. Wu et al^8^ evaluated 8,597 genes in their TWAS and commented that several highly implicated breast cancer susceptibility genes, such as *ESR1, TERT* and *MARS30*, could not be investigated because of poor performance of prediction models. Our study was able to identify these genes as significant at the Bonferroni threshold. Similarly, Feng et al.^7^ investigated only 901 genes in their TWAS.

We identified 29 genes in 11 novel loci, which are at least 1Mb away from any risk variants and not in LD with risk variants reported in previous GWAS. This finding suggests that transcriptome-based association studies are able to discover novel cancer susceptibility signals, extending the capacity of variant-based association studies. These novel genes and loci are plausibly important in breast cancer susceptibility, given evidences from previous studies in other cancers or known cancer pathways. For example, *MAP2K4* in the 17p12 locus, has not been reported to be important in breast cancer susceptibility. We found the predicted expression of *MAP2K4* in multiple tissues, including breast tissue, was positively associated with breast cancer risk. Both colocalization analysis (RCP=0.64) and fine-mapping analysis (PIP=0.893) suggested that *MAP2K4* is a possible causal breast cancer gene and the effect was driven by multiple weak variants. MAP2K4 (a.k.a MKK4) is a member of the MAPK family, which act as an integration point for multiple biochemical signals and are involved in a wide variety of cellular processes such as proliferation, differentiation, transcription regulation, and development. *MAP2K4* has been found to be a metastasis suppressor gene in ovarian carcinoma^25^. Furthermore, *MAP2K4* was identified as a driver gene mutated in both early and metastatic breast cancer ^26,27^. Taken together, it is possible that *MAP2K4* was a breast cancer susceptibility gene in the novel 17p12 locus.

Most of our TWAS-identified genes are located in known GWAS susceptibility loci. Interestingly, we are able to identify possible susceptibility genes in more than half of the known breast cancer GWAS loci. In these scenarios, TWAS revealed possible target genes that risk variants identified in GWAS act on to cause breast cancer. Using eQTL analyses, Guo et al ^28^ inferred 101 target genes in known breast cancer GWAS loci. We re-discovered 51 of these 101 genes in our TWAS.

One interesting GWAS locus is 1p13.3, a gene rich region containing >20 genes within 400kb (Figure 2). Although none of the SNPs in this locus reached GWAS significance in BCAC, this locus was recently reported to be associated with breast cancer in a cross-ancestry study^29^. Of the genes in this region, it is unclear which ones are breast cancer susceptibility genes simply based on GWAS signals. Our TWAS found that the predicted expression of *GSTM1, GSTM2*, and *GSTM4* in multiple tissues, including breast tissue, was inversely associated with breast cancer risk. After adjusting for GWAS index SNPs in conditional analysis, the three genes were no longer significant, suggesting GWAS risk SNPs may be responsible for the observed TWAS signals. Both colocalization analysis (RCP>0.99) and fine-mapping analysis suggested that all three genes are possible breast cancer candidate genes. *GSTM1, GSTM2*, and *GSTM4* are members of the glutathione S-transferase multigene family, which can detoxify xenobiotics, including carcinogenic compounds and thus were proposed as cancer susceptibility genes,^30^ and the *GSTM1* null genotype has been associated with risk of several other cancers^31-37^. Therefore, there exists evidence that all three genes are possible cancer suppressors responsible for GWAS signals in 1p13.3 through carcinogen metabolism.

Another interesting example is the *TNFSF10* gene in 3q26.21 (Figure 2). This gene was first implicated in a GWAS of African ancestry population for estrogen receptor (ER)-negative breast cancer^38^, and was later implicated in a GWAS of European ancestry population for overall breast cancer^3^; however, different risk SNPs were reported between the two studies. In the present study, we found high expression of *TNFSF10* in several tissues, including breast, was associated with lower risk of breast cancer. This gene is the only significant gene in the 3q26.21 locus; it was not significant in our conditional analysis after adjusting for index SNPs. It had a high RCP (0.67) in colocalization analysis. A recently published study deleted the *TNFSF10* gene from TNBC cells using CRISPR-Cas9 technology and stimulated cells with either poly (I:C), a synthetic analogue of double-stranded RNA virus, or IFN-β, and found that depletion of TNFSF10 clearly reduced poly (I:C)-induced or IFN-β-induced apoptosis^39^. Furthermore, the researcher edited rs13074711, the risk variant reported in previous GWAS^38^, using CRISPR-Cas9 technology, and found it altered *TNFSF10* expression and IFN-β-induced apoptosis ^39^. Taken together, there is strong evidence that *TNFSF10* is a possible cancer suppressor responsible for GWAS signal in 3q26.21 through immune defense mechanisms.

Determining causality of TWAS-identified genes remains challenging because these genes may be associated with disease phenotypes through their correlation with disease causal gene(s) in the same LD region. Based on gene-based fine-mapping and colocalization methods, we proposed 58 genes in 47 loci to have a high probability of being causal genes. Still, these genes need to be investigated in future functional experiments. Guo et al ^28^ used luciferase reporter assays to study functional target genes, and they found a significant difference between alternative and reference alleles in promoter activity for five genes (*DCLRE1B, SSBP4, MRPS30, ATG10*, and *PAX9*) but failed to show functional activity for the gene *ARRDC3*. These findings are consistent with ours: We found all five genes were TWAS significant and four of them (*DCLRE1B, SSBP4, ATG10*, and *PAX9)* are in our proposed list of causal genes (Table 3). We did not find *ARRDC3* to be TWAS significant.

The current study has several limitations. First, although the joint TWAS identified more genes than single-tissue TWAS, it may generate more false positive hits because it utilizes other tissues not relevant to phenotype of interest ^40^. However, the target tissue for cancer development might not be distinct, and gene expression across multiple tissues could be partially correlated ^10,11^. We also observed moderate consistency between results of single tissue TWAS. For instance, breast tissue is presumably the target tissue for breast cancer, but gene expression in liver might better reflect carcinogen metabolism. Fortunately, the ACAT method used in our joint TWAS analysis calculates a weighted average of p-values from multiple tissues and is relatively conservative in identifying significant genes. Strikingly, the top tissues in which the joint TWAS-identified genes were upregulated were all female tissues (breast, uterus, ovary and vagina), suggesting that our joint TWAS method was able to automatically prioritize target tissues. One focus of our future method research is to develop more efficient methods to combine TWAS signals across tissues by effectively accounting for the correlation of the signals across tissues or giving high weights to potential target tissues.

Second, the current study only analyzed data from European ancestry and focused on overall breast cancer risk. Breast cancer is a heterogeneous disease consisting of several molecular subtypes. The genetic architecture of estrogen-receptor (ER) negative breast cancer may be different from the estrogen-receptor positive subtype. In the BCAC consortium, 76% patients had ER-positive breast cancer^3^, so the current study may mainly identify genes for susceptibility of ER-positive breast cancer. Future TWAS studies that focus on ER-negative breast cancer or in other racial/ethnic populations are highly desirable. Lastly, the current study only examined overall expression of genes but did not consider the effect of RNA splicing on disease etiology. Li et al.^41^ reported that RNA splicing is another primary link between genetic variation and complex diseases. Therefore, TWAS evaluating associations of genetically predicted splicing with breast cancer have great promise for identifying novel putative candidate disease genes. We are currently working on a splicing-based TWAS of breast cancer.

In conclusion, our joint TWAS identified more than 300 breast cancer genes for further functional investigation. Our approach has discovered new susceptibility loci and mapped out candidate genes in multiple known susceptibility loci. Future studies in diverse populations and with a focus on homogenous phenotypes of breast cancer using innovative TWAS methodology are warranted. There is potential to map out most candidate genes in GWAS loci of breast cancer, the most common malignancy affecting women across the world.

## URLs

PrediXcan GTEx v8 MASHR models, https://predictdb.org/; BCAC summary statistics, https://bcac.ccge.medschl.cam.ac.uk/bcacdata/oncoarray/oncoarray-and-combined-summary-result; UK Biobank, http://ukbiobank.ac.uk; FUMA software, http://fuma.ctglab.nl; Plink 2.0, https://www.cog-genomics.org/plink/2.0/

## Methods

### GWAS summary statistics from the BCAC on women of European ancestry

In our meta-analysis, we used the summary statistics data from the GWAS of breast cancer in 122,977 cases and 105,974 controls of European ancestry from the BCAC. The details of the BCAC have been described previously ^3,42^. Briefly, the BCAC included: 1) 61,282 female cases with breast cancer and 45,494 female controls of European ancestry that were genotyped using the OncoArray including 570,000 SNPs; 2) 46,785 breast cancer cases and 42,892 controls of European ancestry from Collaborative Oncological Gene-environment Study (iCOGS) that were genotyped using a custom Illumina iSelect genotyping array containing ∼211,155 variants; and 3) 11 other breast cancer genome-wide association studies (GWAS; 14,910 cases and 17,588 controls). Genotype data from iCOGS, OncoArray, and GWAS were imputed using the October 2014 release of the 1000 Genomes Project data as a reference. Genetic association results for breast cancer risk were combined using inverse-variance fixed-effect meta-analyses ^3^.

### GWAS analysis using data from UK Biobank

The UK Biobank project recruited approximately 500,000 participants, ages 40 to 69, between 2007 and 2010, across 22 study centers in the United Kingdom. The project collected detailed demographic, lifestyle, and disease histories at baseline, as well as disease occurrences through prospective follow-up and database linkages^13^. Whole-genome genotyping was conducted using UK Biobank Axiom Arrays for 488,377 participants, and imputation was performed using Haplotype Reference Consortium and 1000 Genomes phase 3 as reference panels to obtain >90 million genetic markers^13^. In this study, we selected female individuals with both phenotypic and genotypic data available. Unrelated individuals with European ancestry were selected using principal component analysis. We further filtered out samples with genotyping call rate <5%. After these exclusions, the analysis included 10,534 breast cancer cases and 185,116 controls. We performed GWAS analysis using logistic regression adjusting for age (age at diagnosis for cases and age at last date of follow-up for controls) and top ten eigenvectors from principal component analysis with software package Plink 2.0^43^.

### Gene expression prediction models

Gene expression prediction models were built with the genotype and RNA-seq data in 49 tissues of European ancestry from the GTEx project (v8)^12^. Specifically, building prediction models for a gene includes the following steps: 1) Across all tissues, cis-eQTLs were discovered with a false discovery rate of 5% per tissue. Only genes with cis-eQTLs were selected. 2) Fine mapping was performed in each tissue in the corresponding cis gene region by the dap-g method ^20,44^ to select variants with minor allele frequency > 0.01 and posterior inclusion probabilities (PIPs) > 0.01. Then in each credible set, only the variant with top PIP was kept. For the 49 tissues, a union of selected variants across 49 tissues were obtained and LD pruning was applied to the union of variants to remove redundant variants. 3) The MASH method was applied to the marginal eQTL effects across the 49 tissues at the union of variants to jointly estimate effects of eQTLs, allowing sparse effects (that is, with many zero effects) and accounting for correlation among non-zero effects in different tissues ^14^. 4) The predicted expression of the gene in each tissue was calculated as the linear combination of genotypes multiplying by their estimated effect sizes. In this study, the prediction models for two tissues (prostate and testis) were removed as they were not relevant to breast cancer in women.

### Summary statistic-based imputation

For variants included in the GTEx prediction models but not in the GWAS summary statistics, we imputed z-scores with the method ImpG-Summary ^45^. The ImpG-Summary method assumes under null hypothesis, the vector ***Z*** of z-scores at all SNPs in a locus is approximately distributed as a Gaussian distribution, ***Z***∼ *N*(**0, ∑**) with **∑** being the correlation matrix among all pairs of SNPs induced by LD and estimates posterior mean of z-scores at unobserved SNPs. We used the GWAS summary statistics and correlation matrix estimated by using the genotype data in the GTEx samples as input of the ImpG-Summary method.

### Joint TWAS across multiple tissues

The joint TWAS analysis includes two steps: 1) performing traditional TWAS analysis in each of the 47 tissues by the software S-Predixcan ^16^ to obtain the p-values *p*_*k*_ (*k* = 1, …, 47), and 2) constructing test statistic by ACAT method ^17^ that combined p-values for each gene from the single tissue TWAS analyses across the 47 tissues. Specifically, the ACAT test statistic is 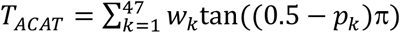, where *w*_*k*_ are nonnegative weights. We used *w*_*k*_ = 1/47. The p-value of the ACAT test statistic is approximated by 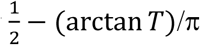.

### Conditional joint TWAS

To test if the signals at the 380 genes by the joint TWAS are independent of a set of published GWAS index SNPs (Supplementary Table S6), we performed TWAS conditional on these index SNPs that were genome-wide significant (p<5×10^−8^). At each gene, we considered two sets of SNPs: the target set of SNPs used for predicting gene expression and the conditioning set of significant index SNPs from published GWAS within ±2 Mb of the transcription start or stop sites of the gene. For the target set of SNPs, we calculated adjusted effects (beta) on breast cancer risk and their variances, after conditioning on the index SNPs using the conditional and joint multiple-SNP (COJO) analysis method of Yang et al ^46^. We then ran S-Predixcan ^16^ on these conditional summary statistics in single tissues and performed the joint TWAS analysis that combines p-values from the single tissue analyses using the ACAT method^17^.

### Colocalization analysis

For the 378 genes identified by the joint TWAS, we calculated RCPs by the method ENLOC ^20^. ENLOC divides the genome into roughly independent LD blocks using the approach described in Berisa & Pickrell ^47^. For a gene located in a specific LD block, ENLOC calculates the colocalization probability of causal GWAS hits and causal eQTLs in the LD block. To calculate RCP for a gene in an LD block, we used the GTEx (v8) eQTLs for the gene and the meta-analysis GWAS summary statistics in the LD block.

### Gene-based fine mapping

We performed a gene-based statistical fine-mapping over the gene– trait association signals from TWAS using the software package FOCUS (fine-mapping of causal gene sets) ^19^. For a LD block, FOCUS estimate sets of genes that contain the causal genes at a predefined confidence level ρ (that is, ρ-credible gene sets; for example, ρ = 90%). FOCUS also computes the marginal PIP for each gene in the region to be causal given the observed TWAS statistics. FOCUS accounts for the correlation structure induced by LD and prediction weights used in the TWAS and controls for certain pleiotropic effects. FOCUS takes as input GWAS summary data, expression prediction weights, and LD among all SNPs in the risk region. FOCUS assumes a single target causal tissue. When the expression prediction model at a gene in the target causal tissue is unavailable, alternative tissues with correlated expression levels are used as a proxy. We used 47 tissues and related expression prediction weights from the GTEx v8 and assigned the breast tissue as the target causal tissue.

### Gene Set Enrichment and Functional Annotation

For the set of 380 significant genes identified by the joint TWAS, we conducted enrichment of protein-coding and lncRNA genes against gene sets from multiple biological pathways, functional categories, and databases by the FUMA package ^21^. Specifically, we used the GENE2FUNC module of FUMA and specified 33,527 protein-coding and lncRNA genes as the background genes for enrichment testing. Multiple testing correction was performed per data source of tested gene sets (e.g., canonical pathways, GWAScatalog categories) using Bonferroni adjustment. We reported pathways/categories with adjusted P-value ≤ 0.05 and at least 2 genes that overlapped with the gene set of interest.

## Data Availability

All data produced in the present study are available upon reasonable request to the authors

https://predictdb.org/

https://bcac.ccge.medschl.cam.ac.uk/bcacdata/oncoarray/oncoarray-and-combined-summary-result

http://ukbiobank.ac.uk

http://fuma.ctglab.nl

https://www.cog-genomics.org/plink/2.0/

## Acknowledgements

This work was supported by the National Cancer Institute (R01-CA242929, R01-CA228198), Breast Cancer Research Foundation (BCRF-21-071), and the NIDDK (P30 DK20595).

For BCAC data, the breast cancer genome-wide association analyses were supported by the Government of Canada through Genome Canada and the Canadian Institutes of Health Research, the ‘Ministère de l’Économie, de la Science et de l’Innovation du Québec’ through Genome Québec and grant PSR-SIIRI-701, The National Institutes of Health (U19 CA148065, X01HG007492), Cancer Research UK (C1287/A10118, C1287/A16563, C1287/A10710) and The European Union (HEALTH-F2-2009-223175 and H2020 633784 and 634935). All studies and funders are listed in Michailidou et al (Nature, 2017). This research has been conducted using the UK Biobank Resource under the Application Number 49564. The authors thank the participants, investigators, and staff of the UK Biobank for providing them with the resources to pursue this research. We thank Sarah Sumner for help editing the paper.

---

**Supplementary Figure S1**. Manhattan plots of joint TWAS and GWAS

**Supplementary Figure S2**. Scatter plot of Z scores from tissue-specific TWAS in BCAC and UKB datasets

**Supplementary Figure S3**. Differential analysis of expression of the joint TWAS-identified genes in GTEx v8 shows tissue specificity

**Supplementary Table S1**. The 378 genes identified by joint TWAS.

**Supplementary Table S2**. The 141 genes identified by breast tissue-based TWAS

**Supplementary Table S3**. Results of single tissue based-TWAS analysis of 380 genes by two databases and across tissue types

**Supplementary Table S4**. List of genes identified in previous TWAS of breast cancer.

**Supplementary Table S5**. List of implicated genes in previous GWAS of breast cancer, curated by GWAS Catalog.

**Supplementary Table S6**. List of known loci and index SNPs reported in previous breast cancer GWAS, curated by GWAS Catalog.

**Supplementary Table S7**. Bonferroni significant gene sets in the enrichment analysis using FUMA

## Notes

### Competing Interest Statement

The authors have declared no competing interest.

### Author Declarations

BCAC summary statistics, https://bcac.ccge.medschl.cam.ac.uk/bcacdata/oncoarray/oncoarray-and-combined-summary-result; UK Biobank, http://ukbiobank.ac.uk;

